# Quality of Care at childbirth during the COVID-19 pandemic: findings of the IMAgiNE EURO study in Belgium

**DOI:** 10.1101/2024.03.25.24304838

**Authors:** Anna Galle, Silke D’Hauwers, Helga Berghman, Nele Vaerewijck, Emanuelle Pessa Valente, Ilaria Mariani, Arianna Bomben, Stefano delle Vedove, Marza Lazzerini, the IMAgiNE EURO Study Group

**Affiliations:** International Centre for Reproductive Health, Department of Public Health and Primary Care, Ghent University, Ghent, Belgium.; Faculty of Medicine and Health Sciences, Department of Public Health and Primary Care, University Centre for Nursing and Midwifery, Ghent University, Ghent, Belgium; WHO Collaborating Centre for Maternal and Child Health, Institute for Maternal and Child Health IRCCS Burlo Garofolo, Trieste, Italy; Maternal Adolescent Reproductive and Child Health Care Centre, London School of Hygiene & Tropical Medicine London UK

**Keywords:** Quality of Care, Women Care, Newborn Care, Childbirth, Midwifery, COVID 19

## Abstract

**Objectives:** To examine quality of maternal and newborn care (QMNC) around childbirth in facilities in Belgium during the COVID-19 pandemic and trends over time.

**Design:** A cross-sectional observational study.

**Setting:** Data of the IMAgiNE EURO study in Belgium. Participants

Women giving birth in a Belgian facility from March 1, 2020, to May 1, 2023, responded a validated online questionnaire based on 40 WHO standards-based quality measures organised in four domains: provision of care, experience of care, availability of resources, and organizational changes related to COVID-19.

**Primary and secondary outcome measures:** Quantile regression analysis was performed to assess predictors of QMNC; trends over time were tested with the Mann-Kendall test.

**Results:** 897 women were included in the analysis, 67%(n=601) with spontaneous vaginal birth, 13.3%(n=119) with instrumental vaginal birth and 19.7%(n=177) with cesarean section. We found high QMNC scores but also specific gaps in all domains of QMNC. On provision of care, 21.0%(n=166) of women who experienced labor and 14.7%(n=26) of women with a cesarean reported inadequate pain relief; 64.7%(n=74) of women with an instrumental birth reported fundal pressure and 72.3% (n=86) reported that forceps or vacuum cup was used without their consent. On experience of care, 31.1%(n=279) reported unclear communication, 32.9%(n=295) reported that they were not involved in choices,11.5%(n=104) stated not being treated with dignity and 8.1%(n=73) experienced abuse. Related to resources, almost half of the women reported an inadequate number of healthcare professionals (46.2%, n=414). The multivariable analyses showed significantly lower QMNC scores for women with an instrumental vaginal birth. Over time there was a significant increase in QMNC score for ‘experience of care’ and ‘key organizational changes due to COVID-19’.

**Conclusions and relevance:** Although overall QMNC scores were high, findings also suggest gaps in QMNC. Underlying causes of these gaps should be explored to design appropriate interventions and policies.

## BACKGROUND

Childbirth should be a positive experience, ensuring women and their babies reach their full potential for health and wellbeing (1). When analyzing quality of maternity care worldwide, two extreme situations have been described: too little, too late (TLTL) and too much, too soon (TMTS) (2). While TLTL identifies care with inadequate resources, below evidence-based standards, or care withheld or unavailable until too late to help, TMTS identifies care characterized by over-medicalization, including the use of non-evidence-based interventions, or interventions not appropriate for the case (2). Typical examples of overused interventions during childbirth are caesarean sections, inductions or augmentation of labour, episiotomies, and fundal pressure. Both TLTL and TMTS are costly for health systems and can be dangerous for women and newborns ((3–5). In addition, the literature indicates that women, both in low- and high-income countries (HICs) are often not adequately informed and are minimally involved in decision-making prior to conducting these interventions during childbirth ((5–8). Also, other aspects of experience of care such as privacy, quality of communication, respect and dignity have been described as substandard both in low and HICs (9,10).

In Belgium, the quality of maternal and newborn health has been mainly explored focusing on clinical outcomes and the provision of care(11,12). Similar to neighboring European countries, maternal and newborn health outcomes (such mortality and morbidity) are among the best in the world (11,13,14). Nevertheless, reports also show high rates of interventions (such as cesarean sections and episiotomies) with a high variation between hospitals (11). This unexplained variation suggests that interventions are not always performed based on evidence and might be the result of organizational policies and health providers preferences (11). One pre-pandemic study also showed women often experience a lack of involvement in the decision-making process during childbirth in Belgium, negatively affecting experience of care (15). However, more research using validated instruments is needed to capture both experience and provision of childbirth care in Belgium.

On the 11th of March 2020, the WHO declared the COVID-19 pandemic as a public health emergency of international concern; which was declared as ended on May 5, 2023. Globally, studies have shown that COVID–19 negatively impacted the provision and experience of maternal and newborn healthcare, especially in the first year of the pandemic (16–18). Rapidly implemented measures (such as stringent lockdown measures, curfews, isolation of suspected and confirmed cases) to control the pandemic negatively affected the availability, utilization and quality of essential maternal and newborn health services (17). A systematic review showed maternal and fetal outcomes worsened globally during the COVID-19 pandemic, with an increase in maternal deaths, stillbirth, ruptured ectopic pregnancies, and maternal depression (19,20). However, outcomes show considerable disparity between different settings within and across countries (18–20) and changes over time are yet to be explored.

IMAgiNE EURO is a multicountry project that started at the onset of the COVID 19 pandemic, exploring through online surveys the perspective of women and health care providers on the Quality of Maternal and Newborn Care (QMNC) at childbirth in hospital settings. Two validated questionnaires were developed for this project, containing 80 prioritized WHO Quality measures (out of the of more than 300 suggested by the WHO Standards for improving the quality of maternal and newborn care)(21). This paper presents detailed survey findings on QMNC and trends over time, from the perspective of women who gave birth in Belgium during the COVID-19 pandemic, between March 2020 and May 2023.

## METHODS

A cross-sectional observational study was conducted and reported according to the STROBE Strengthening the Reporting of Observational Studies in Epidemiology guidelines and the checklist can be found in supplementary materials (Supplementary Material 1) (22). As first aim overall quality of care was explored according to the WHO Quality of Care Measures. As a secondary aim, trends over time were analyzed for the different subdomains.

### Participants

Only women who gave birth in a Belgian facility between March 2020 and May 2023 were included in this study, corresponding to the period the pandemic was officially declared by WHO (23). Women needed to be 18 years or older to be eligible for participation. Women who gave birth multiple times during the described period could fill in the questionnaire for each childbirth. Women were able to select their preferred language from 28 languages available for completing the questionnaire and participated by actively clicking on the link or scanning the QR code to access the questionnaire.

### Data collection

We used a structured online questionnaire to collect data, recorded with Research Electronic Data Capture (REDCap 8.5.21) via a centralized platform (24). The process of questionnaire development, validation and previous use has been reported elsewhere (25,26) and the study was registered at the U.S. National Library of Medicine under NCT04847336. The questionnaire for women included 40 questions on one key indicator each, equally distributed in four domains: the three domains of the WHO Quality measures (21), namely provision of care, experience of care and availability of human and physical resources, plus an additional domain on key organizational changes related to the COVID-19 pandemic (26).

Two versions of the questionnaire were available, one tailored for women who experienced labor and one for women who did not (e.g., women with a planned cesarean section). Each included the 40 WHO standard-based prioritized quality measure with 34 measures in common. Labor was defined according to NICE guidelines (27). Questions on individual characteristics of the participants (e.g., socioeconomic background, parity) were included. As reported elsewhere (26) 40 indicators contributed to a composite QMNC index, ranging from 0 to 100 for each of the four domains, for a total score ranging from 0 to 400 points, and higher scores indicating higher adherence to the WHO Standards (28). The online questionnaire was disseminated by social media (Facebook and X) and by distributing leaflets in maternity wards, postnatal clinics and creches. Dissemination materials were available in Dutch, French and English.

### Data analysis

#### Sample size

A minimum required sample size of 300 women for each country was calculated, based on preliminary data from other studies on the hypothesis of an average QMNC Index (our primary outcome and dependent variable) of 75% +/-7.5% (300 +/-30 points, out of 400) and confidence level of 99.5%. Women were included when they had less than 20% cases missing on the 40 Quality measures and all questions on the individual characteristics. For the primary aim, we calculated absolute frequencies and percentages for sociodemographic variables and for each of the 40 key quality measures. The QMNC index was presented as median and interquartile range (IQR) because not normally distributed (Shapiro-Wilk test for normality P<0.05).

In addition, we performed a multivariable quantile regression analysis with the QMNC index as the dependent variable and with trimester, maternal age, parity, maternal education, type of facility, mode of birth and presence of an OB/GYN directly assisting childbirth as independent variables. Quantile regression was chosen instead of linear regression since the QMNC index was not normally distributed and owing to evidence of heteroskedasticity ((29). We conducted a multivariable quantile regression with robust standard errors (SEs) and we modeled the median, the 0.25th and 0.75th quantile, given statistical evidence of heteroskedasticity for parity, mode of birth, place of birth of the woman (Breusch-Pagan/Cook-Weisberg test P < 0.05, H0: homoskedasticity). The categories with the highest frequency were used as reference. For our secondary aim, we assessed the hypothesis that the QMNC index improved over time during the pandemic period (30). We first evaluated time trends by trimester for total QMNC index and subsequently for the QMNC index by domain. Time trends were tested with the Mann-Kendall test. All the tests were two-tailed and a p-value <0.05 was considered statistically significant. Statistical analyses were performed using Stata version 14 (Stata Corporation) and R version 4.1.1 (31).

### Ethical aspects

The international study was approved by the Institutional Review Board of the coordinating center: the IRCCS “Burlo Garofolo” Trieste (IRB-BURLO 05/2020 15.07.2020) and the Commision Medical Ethics UZ Ghent (THE-2023-0075). The study was conducted according to General Data Protection Regulation requirements. Participation in the online survey was voluntary and anonymous. Prior to participation, women were informed of the objectives and methods of the study, including their rights in declining participation, and each participant provided consent before responding to the questionnaire. Anonymity in data collection during the survey phase was ensured by not collecting any information that could disclose participant identity, such as facility of birth or day of birth of the woman. Data transmission and storage were secured by encryption.

## RESULTS

### Sociodemographic characteristics

Of 74 026 women accessing the online questionnaire in all participating countries, 52 632 women fulfilled the inclusion criteria, and responses from 897 women giving birth in Belgium were analyzed after data cleaning (Figure 1). The Dutch questionnaire was chosen by 83.8% (n=752) of women, the French questionnaire was chosen by 12.2% (n=109) of women, 1.7% (n=15) chose the English, and the rest opted for one of the other available languages (Table 1).

**Figure 1.**
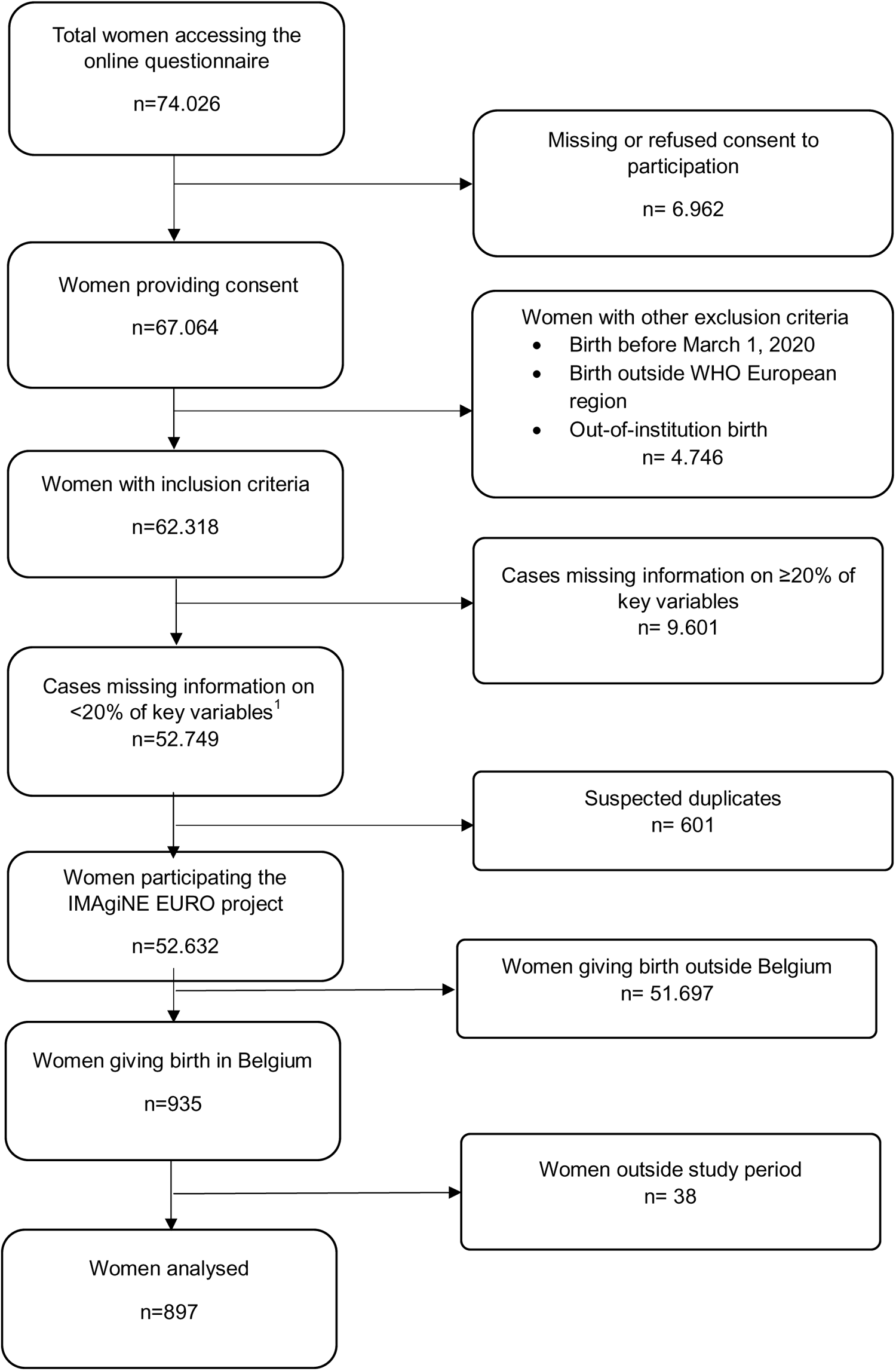
Flowchart of the study sample of women.

Overall, most women (93.7%, n=841) were aged between 25 and 39 years and 54.2% (n=486) of women had university degree or higher. More than half of the women (61.2%, n=549) were primiparous (Table 1). Frequencies of spontaneous vaginal birth (SVB) and instrumental vaginal birth (IVB) were 67.0% (n=601) and 13.3% (n=119), respectively, while frequencies for cesarean during labor, elective, and emergency cesarean before labor were 8% (n=72), 3.1% (n=28), and 8.6% (n=77), respectively. Most women gave birth in a public hospital (89.1%, n=799). Almost all births were assisted by a midwife or nurse (96.8%, n=868) and 87.2% (n=782) of births were assisted by an obstetrics or gynecology doctor. From all women, 11.4% (n=102) had a newborn admitted to the Neonatal Intensive Care Unit (NICU), 1.6% (n=14) had multiple births and 0.7 % (n=6) had a stillbirth.

**Table 1.** Characteristics of responders (N=897)

|  | n | % |
| --- | --- | --- |
| **Year of birth** |  |  |
| 2020 | 313 | 34.9 |
| 2021 | 311 | 34.7 |
| 2022 | 178 | 19.8 |
| 2023 | 95 | 10.6 |
| **Mother giving birth in the same country where she was born** |  |  |
| Yes | 816 | 91.0 |
| No | 81 | 9.0 |
| **Age range (years)** |  |  |
| 18-24 | 21 | 2.3 |
| 25-30 | 298 | 33.2 |
| 31-35 | 436 | 48.6 |
| 36-39 | 107 | 11.9 |
| ≥40 | 35 | 3.9 |
| **Educational level <sup>1</sup>** |  |  |
| None | 0 | 0.0 |
| Elementary school | 6 | 0.7 |
| Junior high school | 94 | 10.5 |
| High school | 311 | 34.7 |
| University degree | 194 | 21.6 |
| Postgraduate degree / Master /Doctorate or higher | 292 | 32.6 |
| **Birth mode** |  |  |
| spontaneous vaginal birth | 601 | 67.0 |
| Instrumental vaginal birth | 119 | 13.3 |
| Caesarian section | 177 | 19.7 |
| **Parity** |  |  |
| 1 | 549 | 61.2 |
| >1 | 348 | 38.8 |
| **Type of hospital** |  |  |
| Public | 799 | 89.1 |
| Private | 98 | 10.9 |
| **Type of healthcare providers who directly assisted birth** |  |  |
| Midwife or nurse | 868 | 96.8 |
| A student (i.g. before graduation) | 302 | 33.7 |
| Obstetrics registrar / medical resident (under post-graduation training) | 285 | 31.8 |
| Obstetrics and gynecology doctor | 782 | 87.2 |
| I don't know (healthcare providers did not introduce themselves) | 33 | 3.7 |
| Other | 64 | 7.1 |
| **Language in which questionnaire was filled** |  |  |
| Dutch | 752 | 83.8 |
| French | 109 | 12.2 |
| English | 15 | 1.7 |
| Polish | 5 | 0.6 |
| German | 3 | 0.3 |
| Latvian | 3 | 0.3 |
| Swedish | 3 | 0.3 |
| Croatian | 2 | 0.2 |
| Greek | 2 | 0.2 |
| Portuguese | 2 | 0.2 |
| Italian | 1 | 0.1 |
| **Other** |  |  |
| Newborn admission to NICU | 102 | 11.4 |
| Maternal admission to ICU | 6 | 0.7 |
| Multiple births | 14 | 1.6 |
| Stillbirth | 6 | 0.7 |

<sup>1</sup>Wording on education levels agreed among partners during the Delphi; questionnaire translated and back translated according to Wild et al. (2005). Abbreviations: NICU =Neonatal Intensive Care Unit

### WHO-based quality measures

Key results for the domain of provision of care (Table 2) were as follows: 21.0% (n=166) of women who experienced labor and 14.7% (n=26) of women who underwent a cesarean reported inadequate pain relief; 64.7% (n=77) of women with an instrumental vaginal birth reported fundal pressure during childbirth; 35.8% (n=215) of women with spontaneous vaginal birth had an episiotomy; 4.6% (n=41) did not experience skin-to-skin contact with their newborn; 12.5% (n=112) reported no early breastfeeding; 10.9% (n=98) were not exclusively breastfeeding at discharge, and 18.6% (n=167) reported inadequate breastfeeding support. One in three women reported they did not receive immediate attention when needed (29.2%, n=262).

For experience of care, 32.9% (n=295) women reported that they were not involved in choices, 72.3% (n=86) were not asked for consent prior to an IVB. Overall, one in ten women stated that they were not treated with dignity (11.6%, n=104), while 8.1% (n=73) were exposed to physical, verbal, or emotional abuse. Nearly one in four women (23.9%, n=189) reported no freedom of movement during labor and 15.9% (n=143) reported a lack of privacy while almost none (1.3%, n=12) reported they performed informal payments. One in three women (31.1%, n=279) mentioned that the communication with healthcare professionals was unclear or ineffective.

For availability of human and physical resources, about half of women (46.2%, n=414) observed that staff were inadequate in number, while around half of women reported they received inadequate information on maternal and newborn danger signs (43.9%, n=394 and 55.4%, n=497, respectively). Room comfort, cleaning, and number of women per room were rated as “inadequate” by 25.9% (n=232), 18.7% (n=168), and 8.9% (n=80) of women respectively, while 24.4% (n=219) respondents judged staff professionalism as inadequate.

For reorganizational changes due to COVID-19, around one in three women (30.9%, n=277) reported that COVID-19 had led to a reduction in QMNC and a high percentage of women reported difficulties in attending routine antenatal checks and experienced barriers in accessing the facility (95.1%, n=853 and 97.5%, n=875, respectively). Regarding staff, 7.4% (n=66) women noted that healthcare personnel were not always using personal protective equipment, while for one in four women (24%, n=215) the communication did not contain their stress related to COVID-19-required procedures. Overall 17.6% (n=158) rated the info graphics as inadequate or noted a lack of handwashing stations (3.8%, n=34).

**Table 2:** Results for WHO standards-based quality measures.

|  | Women experiencing labor |  | Women not experiencing labor |  | Overall (women experiencing labor and women not experiencing labor) |  |
| --- | --- | --- | --- | --- | --- | --- |
|  | N=792 |  | N=105 |  | N=897 |  |
|  | N | % | N | % | N | % |
| Provision of care |  |  |  |  |  |  |
| 1. No pain relief during labor | 166 | 21 | - | - | - | - |
| 2. Mode of birth |  |  |  |  | - | - |
| 2a. SVB | 601 | 75.9 | - | - | - | - |
| 2b. IVB | 119 | 15 | - | - | - | - |
| 2c. CS after labor | 72 | 9.1 | - | - | - | - |
| 2d. CS before labor | - | - | 28 | 26.7 | - | - |
| 2e. Elective cesarean | - | - | 77 | 73.3 | - | - |
| 3a. Episiotomy (in SVB) | 215/601 | 35.8 | - | - | - | - |
| 3b. Fundal pressure (in IVB) | 77/119 | 64.7 | - | - | - | - |
| 3c. No pain relief after cesarean | 9/72 | 12.5 | 17 | 16.2 | 26/177 | 14.6 |
|  | N=792 |  | N=105 |  | N=897 |  |
|  | N | % | N | % | N | % |
| 4. No skin-to-skin contact | 23 | 2.9 | 18 | 17.1 | 41 | 4.6 |
| 5. No early breastfeeding | 84 | 10.6 | 28 | 26.7 | 112 | 12.5 |
| 6. Inadequate breastfeeding support | 149 | 18.8 | 18 | 17.1 | 167 | 18.6 |
| 7. No rooming-in | 71 | 9 | 13 | 12.4 | 84 | 9.4 |
| 8. Not allowed to stay with the baby as wished | 40 | 5.1 | 7 | 6.7 | 47 | 5.2 |
| 9. No exclusive breastfeeding at discharge | 83 | 10.5 | 15 | 14.3 | 98 | 10.9 |
| 10. No immediate attention when needed | 230 | 29 | 32 | 30.5 | 262 | 29.2 |
| Experience of care | N | % | N | % |  |  |
| 1a. No freedom of movements during labor | 189 | 23.9 | - | - | - | - |
| 1b. No consent requested for vaginal examination before prelabour cesarean | - | - | 21 | 20 | - | - |
| 2a. No choice of birth position (in SVB) | 276/601 | 45.9 | - | - | - | - |
| 2b. No consent requested (for IVB) | 86/119 | 72.3 | - | - | - | - |
| 2c. No information on newborn (after cesarean) | 16/72 | 22.2 | 16 | 15.2 | 32/172 | 18.1 |
| 3. No clear/effective communication from HCP | 251 | 31.7 | 28 | 26.7 | 279 | 31.1 |
| 4. No involvement in choices | 260 | 32.8 | 35 | 33.3 | 295 | 32.9 |
| 5. Companionship not allowed | 74 | 9.3 | 10 | 9.5 | 84 | 9.4 |
| 6. Not treated with dignity | 88 | 11.1 | 16 | 15.2 | 104 | 11.6 |
| 7. No emotional support | 196 | 24.7 | 29 | 27.6 | 225 | 25.1 |
|  | N=792 |  | N=105 |  | N=897 |  |
|  | N | % | N | % | N | % |
| 8. No privacy | 128 | 16.2 | 15 | 14.3 | 143 | 15.9 |
| 9. Abuse (physical/verbal/emotional) | 62 | 7.8 | 11 | 10.5 | 73 | 8.1 |
| 10. Informal payment | 11 | 1.4 | 1 | 1 | 12 | 1.3 |
| Availability of physical and human resources |  |  |  |  |  |  |
| 1a. No timely care by HCPs at facility arrival | 71 | 9 | 11 | 10.5 | 82 | 9.1 |
| 2. No information on maternal danger signs | 343 | 43.3 | 51 | 48.6 | 394 | 43.9 |
| 3. No information on newborn danger signs | 442 | 55.8 | 55 | 52.4 | 497 | 55.4 |
| 4. Inadequate room comfort and equipment | 203 | 25.6 | 29 | 27.6 | 232 | 25.9 |
| 5. Inadequate number of women per rooms | 69 | 8.7 | 11 | 10.5 | 80 | 8.9 |
| 6. Inadequate room cleaning | 145 | 18.3 | 23 | 21.9 | 168 | 18.7 |
| 7. Inadequate bathroom | 134 | 16.9 | 16 | 15.2 | 150 | 16.7 |
| 8. Inadequate partner visiting hours | 320 | 40.4 | 46 | 43.8 | 366 | 40.8 |
| 9. Inadequate number of HCPs | 364 | 46 | 50 | 47.6 | 414 | 46.2 |
| 10. Inadequate HCP professionalism | 197 | 24.9 | 22 | 21 | 219 | 24.4 |
| Reorganizational changes due to COVID-19 |  |  |  |  |  |  |
| 1. Difficulties in attending routine antenatal Visits | 751 | 94.8 | 102 | 97.1 | 853 | 95.1 |
| 2. Any barriers in accessing the facility | 770 | 97.2 | 105 | 100 | 875 | 97.5 |
| 3. Inadequate info graphics | 137 | 17.3 | 21 | 20 | 158 | 17.6 |
|  | N=792 |  | N=105 |  | N=897 |  |
|  | N | % | N | % | N | % |
| 4. Inadequate ward reorganization | 116 | 14.6 | 17 | 16.2 | 133 | 18.7 |
| 5. Inadequate room reorganization | 109 | 13.8 | 13 | 12.4 | 122 | 13.6 |
| 6. Lacking one functioning accessible handwashing station | 28 | 3.5 | 6 | 5.7 | 34 | 40.8 |
| 7. HCP not always using PPE | 59 | 7.4 | 7 | 6.7 | 66 | 7.4 |
| 8. Insufficient HCP number | 205 | 25.9 | 33 | 31.4 | 238 | 26.5 |
| 9. Communication inadequate to contain COVID | 191 | 24.1 | 24 | 22.9 | 215 | 24.0 |
| 10. Reduction in QMNC due to COVID-19 | 238 | 30.1 | 39 | 37.1 | 277 | 30.9 |

Notes: Indicators identified with letters (eg, 3a, 3b) were tailored to take into account different mode of birth (ie, spontaneous vaginal, instrumental vaginal, and caesarean section). These were calculated on subsamples (e.g., 3a was calculated on spontaneous vaginal births; 3b was calculated on instrumental vaginal births). Indicator 6 in the “reorganizational changes due to COVID-19” domain was defined as: at least one functioning and accessible hand-washing station (near or inside the room where the mother was hospitalized) supplied with water and soap or with disinfectant alcohol solution.
Abbreviations: CS =caesarean section; HCP =health care provider; IVB =instrumental vaginal birth; PPE =personal protective equipment; QMNC =quality maternal and newborn care; SVB =spontaneous vaginal birth

### Predictors of QMNC indexes

Multivariable analysis showed that when adjusting the QMNC index for other variables, only minor differences among groups were observed, except for women who had an instrumental vaginal birth (Table 3). Significantly lower QMNC indexes were reported by women aged above 40 (-12 in the 75^th^ percentile, P=0.027) and women who had an instrumental vaginal birth (−23.1 in the 25th centile, P =0.009; -20.4 in the 50^th^ centile, P<0.001; -20 in the 75^th^ centile, P<0;001). Significantly higher QMNC index was reported on selected centiles for women who were not born in Belgium (+12 in the 75th centile, P=0.004), with a university degree (+9.4 in the 50th centile, P=0.042) or postgraduate degree (+12 in the 50^th^ percentile, P=0.013; +9 in the 75^th^ percentile, P=0.02) and who gave birth in a facility with private offers (+8.7 in the 50^th^ percentile, P=0.046).

**Table 3:** Multivariate analysis predictors QMNC index.

|  | 25th percentile |  | 50th percentile |  | 75th percentile |  |
| --- | --- | --- | --- | --- | --- | --- |
|  | Coefficient<br>(95%CI) | p-value | Coefficient<br>(95%CI)2 | p-value2 | Coefficient<br>(95%CI)3 | p-value3 |
| Study trimester | 1.55 (0.9; 2.2) | 0.019 | 0.93 (0.4; 1.5) | 0.102 | 0.5 (0.1; 0.9) | 0.261 |
| Parity |  |  |  |  |  |  |
| >1 | -7.8 (-13.1; -2.5) | 0.144 | -4.8 (-8.9; -0.7) | 0.239 | 0.5 (-2.6; 3.6) | 0.87 |
| 1 | Ref |  | Ref |  | Ref |  |
| Mother giving birth in the same country where she was born |  |  |  |  |  |  |
| No | 12.2 (3.1; 21.4) | 0.182 | 11.3 (2.3; 20.3) | 0.209 | 12 (7.8; 16.2) | 0.004 |
| Yes | Ref |  | Ref |  | Ref |  |
| Type of facility |  |  |  |  |  |  |
| Private | 12.6 (3.8; 21.3) | 0.151 | 8.7 (4.4; 13.1) | 0.046 | 8.5 (3.8; 13.2) | 0.072 |
| Public | Ref |  | Ref |  | Ref |  |
| Maternal age |  |  |  |  |  |  |
| 18-24 | -7.1 (-39.5; 25.4) | 0.827 | -0.4 (-18.6; 17.8) | 0.984 | -1.5 (-17.5; 14.5) | 0.925 |
| 25-30 | -12.1 (-18.3; -5.8) | 0.055 | -4.6 (-9.6; 0.3) | 0.352 | 1.5 (-2; 5) | 0.669 |
| 31-35 | Ref |  | Ref |  | Ref |  |
| 36-39 | 8.4 (-0.1; 17) | 0.322 | 6.3 (0.6; 12) | 0.267 | 1.5 (-2.2; 5.2) | 0.687 |
| ≥40 | 7.6 (-0.2; 15.4) | 0.333 | -5.7 (-12.9; 1.4) | 0.424 | -12 (-17.4; -6.6) | 0.027 |
| Maternal education |  |  |  |  |  |  |
| Elementary school | 4 (-43.7; 51.6) | 0.934 | -22.8 (-55.2; 9.6) | 0.482 | -3 (-27.2; 21.2) | 0.901 |
| Junior high school | -15.7 (-29; -2.4) | 0.240 | -11.7 (-19.4; -3.9) | 0.132 | -8 (-17; 1) | 0.374 |
| High school | Ref |  | Ref |  | Ref |  |
| University degree | 4.7 (-2.4; 11.7) | 0.510 | 9.4 (4.8; 14.1) | 0.042 | 5 (1.1; 8.9) | 0.203 |
| Postgraduate degree / Master /Doctorate or higher | 10.2 (5; 15.3) | 0.050 | 12 (7.2; 16.9) | 0.013 | 9 (5.1; 12.9) | 0.020 |
| Mode of birth |  |  |  |  |  |  |
| Spontaneous vaginal birth | Ref |  | Ref |  | Ref |  |
| Instrumental vaginal birth | -23.1 (-31.9; -14.3) | 0.009 | -20.4 (-25.2; -15.5) | <0.001 | -20 (-24.5; -15.5) | <0.001 |
|  | Coefficient<br>(95%CI) | p-value | Coefficient<br>(95%CI)2 | p-value2 | Coefficient<br>(95%CI)3 | p-value3 |
| Caesarian section | -12.2 (-20.4; -4.1) | 0.132 | -6.1 (-11.3; -0.9) | 0.24 | -3 (-6.8; 0.8) | 0.432 |
| Presence of an OB/GYN directly assisting childbirth |  |  |  |  |  |  |
| Yes | 17.2 (10.2; 24.2) | 0.014 | 16.1 (8.8; 23.4) | 0.028 | 9 (4.7; 13.3) | 0.039 |
| No | Ref |  | Ref |  | Ref |  |

Abbreviations: OB/GYN=obstetrician/gynecologist

### Trends over time for QMNC indexes

For the QMNC index of experience of care and key organizational changes related to the COVID-19 pandemic, a steady increase over time was observed. Experience of care increased from a median score of 85 points in the first study trimester to 95 points at study end, and key organizational changes related to the COVID-19 pandemic evolved from a median score of 90 points in the first trimester to 95 points at study (trend test *P* < 0.05) (Figure 2). The QMNC indexes in the domains of provision of care and availability of human and physical resources did not show any significant trend over time (see supplementary material 2).

**Figure 2:**
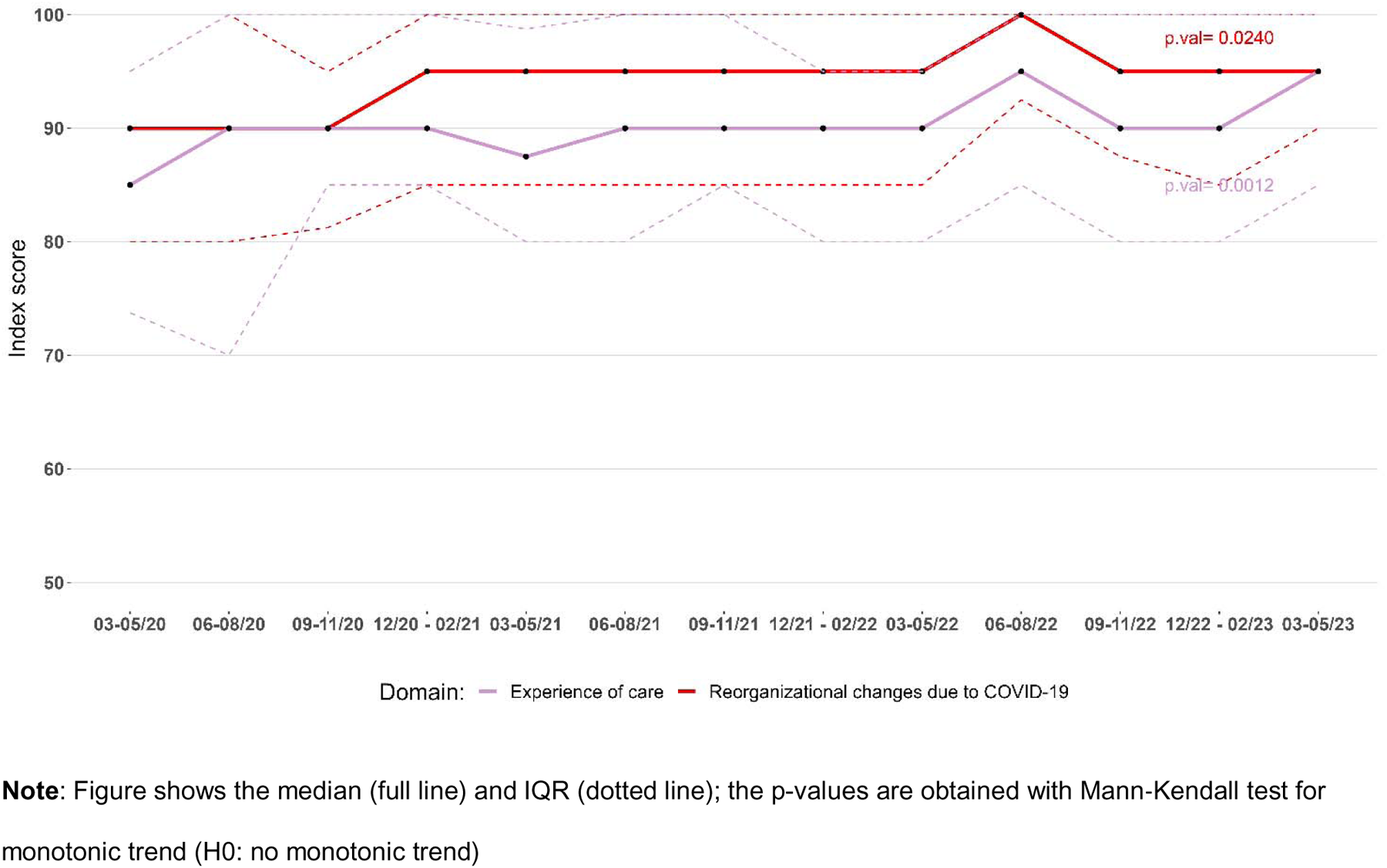
Lineplot showing the Experiences of care and reorganizational changes due to COVID-19 indexes by study trimester.

## DISCUSSION

This is the first study exploring the perceived QMNC care at birth facilities in Belgium during COVID-19, using a set of 40 Quality measures based on WHO Standards. Many of the quality measures explored, such as those related to the high rate of early breastfeeding, skin-to-skin contact, privacy and timely care, suggest high QMNC in Belgium. This is in line with the literature showing high satisfaction with maternity care in Belgium and high rankings of maternal and newborn health outcomes in Europe (14,32–34). However, gaps in QMNC were also reported in our study for each domain: provision of care (e.g. no pain relief after cesarean and inadequate breastfeeding support); experience of care (lack of consent request and involvement in choices); availability of resources (inadequate number of HCPs and HCP professionalism) and reorganizational changes (barriers in accessing the facility and reduction in QMNC due to COVID-19). Reports showed an improved trend over time in the domains of reorganizational changes due to COVID-19 and experience of care suggesting that the impact of COVID-19 was most severe in the first months of the pandemic.

Some study findings are of particular concern. In the domain of provision of care, the results showed that 64.6% of women with an IVB are subjected to fundal pressure. This is surprising, since both WHO and FIGO do not recommend this practice, given its safety is yet unproven ((35,36). Also, national guidelines state that “there are no medically validated indications for the application of fundal pressure. The traumatic experience of patients and their families and the occurrence of rare but serious complications are reasons for discontinuing its use” (37). More research will be needed to explore the underlying reasons for the high rates of fundal pressure in this study and how adherence to national and international guidelines can be improved. Also in the immediate post-partum period, we observed shortcomings in provision of care: a relative high proportion of women with a cesarean did not receive any pain relief and breastfeeding support seem inadequate. While quality of childbirth care is often evaluated by internal audits and registry data, the (immediate) postpartum care is often receiving less attention. Maternity wards might need more close feedback and audit mechanisms of women and their families to improve quality of (immediate) postpartum care and identify specific breaches in their organization (38,39). Especially since there has been a continuing tendency in Belgium to improve efficiency and close smaller maternity units, which might have affected quality of care in a negative way (40,41).

With respect to experience of care, a high proportion of women state involvement in decisions was limited, communication was inadequate, and consent was not requested before performing interventions. Women with an instrumental birth also reported lower QMNC scores. In addition, 11.6% of women reported she did not feel treated with dignity and 8.1% reported a form of abuse. Lack of communication and autonomy during childbirth is a serious concern and should be tackled to improve women’s experience. A recent review showed that communication skills can be enhanced by training and using additional communication tools, however, the importance of an enabling environment cannot be underestimated (42). The health care system in which health providers operate undoubtedly impacts the ability to effectively communicate and respect women’s choices. Enabling factors can include a non-excessive workload (allowing time to communicate), availability of adequate space and resources, and a work atmosphere where teamwork, empathy and good communication are the norm (42). More research into these environmental factors and providers’ perspectives is highly needed in the Belgian context to tackle these shortcomings in childbirth care.

Related to the availability of human resources (inadequate number of HCPs and HCP professionalism) and reorganizational changes (barriers in accessing the facility and reduction in QMNC due to COVID-19) our study shows similar results as neighboring countries. These findings confirm the numerous studies highlighting the indirect effects of COVID 19 on QMNC (43–49). A study from the UK showed how mitigation measures caused social isolation of women and delays in care (46), while global reports show overall higher levels of fear and stress among both health providers and women (43,45).

Our findings also showed that for the domains of experience of care and key organizational changes due to COVID-19 the QMNC indexes increased over time, which can be explained by several factors, including a better organization of care over time, downscaling of restrictions, and better anticipation of women on mitigation measures. Unfortunately, the lack of previous studies investigating comprehensively maternal perceptions of the QMNC (with the same WHO Quality measures used in this study) make it impossible to further assess to which extend the study findings may be associated with the pandemic. The IMAgiNE EURO study will perform other rounds of data collection, which will allow us to explore if quality gaps persist beyond the COVID-19 pandemic.

## LIMITATIONS AND STRENGHTS

Limitations and strengths of the multicounty IMAgiNE EURO survey have been described elsewhere (5). Specific limitations to this study in Belgium are related to the convenience sampling procedure. Our study sample is highly comparable to the overall population (based on national registry data of women giving birth in Belgium) but we observed a slightly higher proportion of younger (<25) and primiparous women (11,12). This difference might be related to the recruitment by social media, which might be more accessible to the younger population. In addition, only data from women was collected in this study. Data from health providers is needed, as well as qualitative data from women, to understand the underlying mechanisms causing the different gaps in quality of care in Belgium.

## CONCLUSION

While the evidence overall suggests high QMNC in Belgium, our findings also highlighted several gaps in care. These gaps include inadequate and/or unclear communication from health care providers, lack of involvement in choices, inadequate staff number, frequent use of fundal pressure and inadequate pain management. These reported gaps in care should be analyzed more in depth from a health system-based perspective to identify underlying causes and design appropriate interventions and policies. We also found that women’s experience of QMNC improved during the study period, but further research is needed to gain knowledge on QMNC beyond the pandemic and trends over time.

## Data Availability

All data produced in the present study are available upon reasonable request to the authors

## DECLARATIONS

### Availability of data and materials

Data are available upon reasonable request to the corresponding author.

### Author contributions

ML conceived the IMAgiNE EURO study, with major inputs from EPV. AG and SD promoted the survey in Belgium. AG wrote a first draft with input from SDV and IM for the methods and data analysis. IM and SDV analyzed the data. AG wrote the final draft, with major input from all authors. All authors approved the final version of the manuscript for submission.

## Acknowledgements

This work was supported by the Ministry of Health, Rome, Italy, in collaboration with the Institute for Maternal and Child Health IRCCS “Burlo Garofolo”, Trieste, Italy. We would like to thank all women who took their time to respond to this survey. We thank our colleagues from the Department of Public Health and Primary Care Ghent University, the involved maternity hospitals, and others who helped in the dissemination of the invitation to participate in the survey. Special thanks to the IMAgiNE EURO study group for their contribution to the development of this project and support for this manuscript.

## Disclaimer

The authors are responsible for the views expressed in this article and do not necessarily represent the views, decisions, or policies of the institutions they are affiliated with.

