## Supplemental Material 2 for "Quality of Care at childbirth during the COVID-19 pandemic: findings of the IMAgiNE EURO study in Belgium"

**Figure 2A: Lineplot showing the scores of QMNC index by trimester**

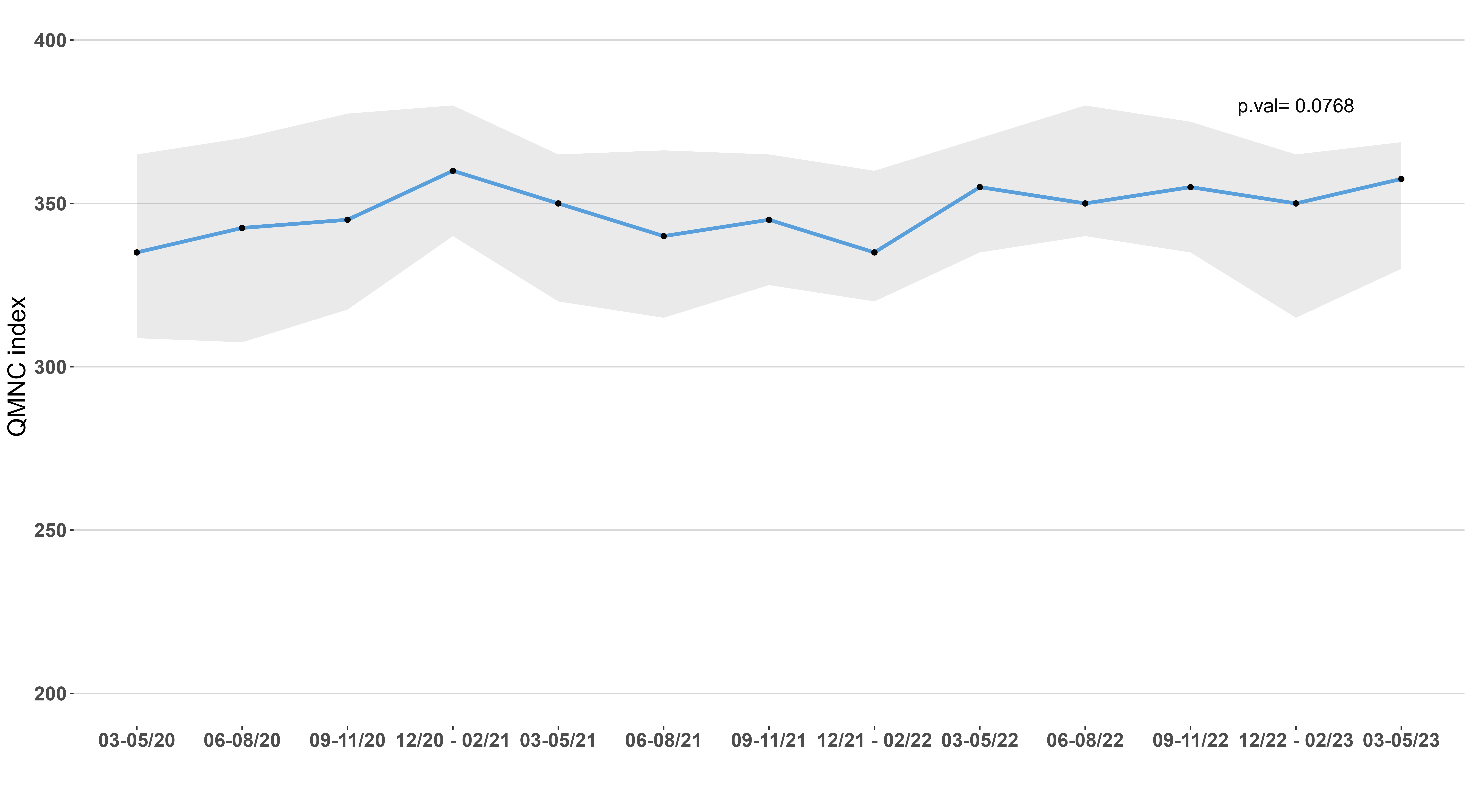

**Note**: the p-value is obtained with Mann‐Kendall test for monotonic trend (H0: no monotonic trend)

**Figure 2B: Lineplot showing the scores of Provision of care index by trimester**
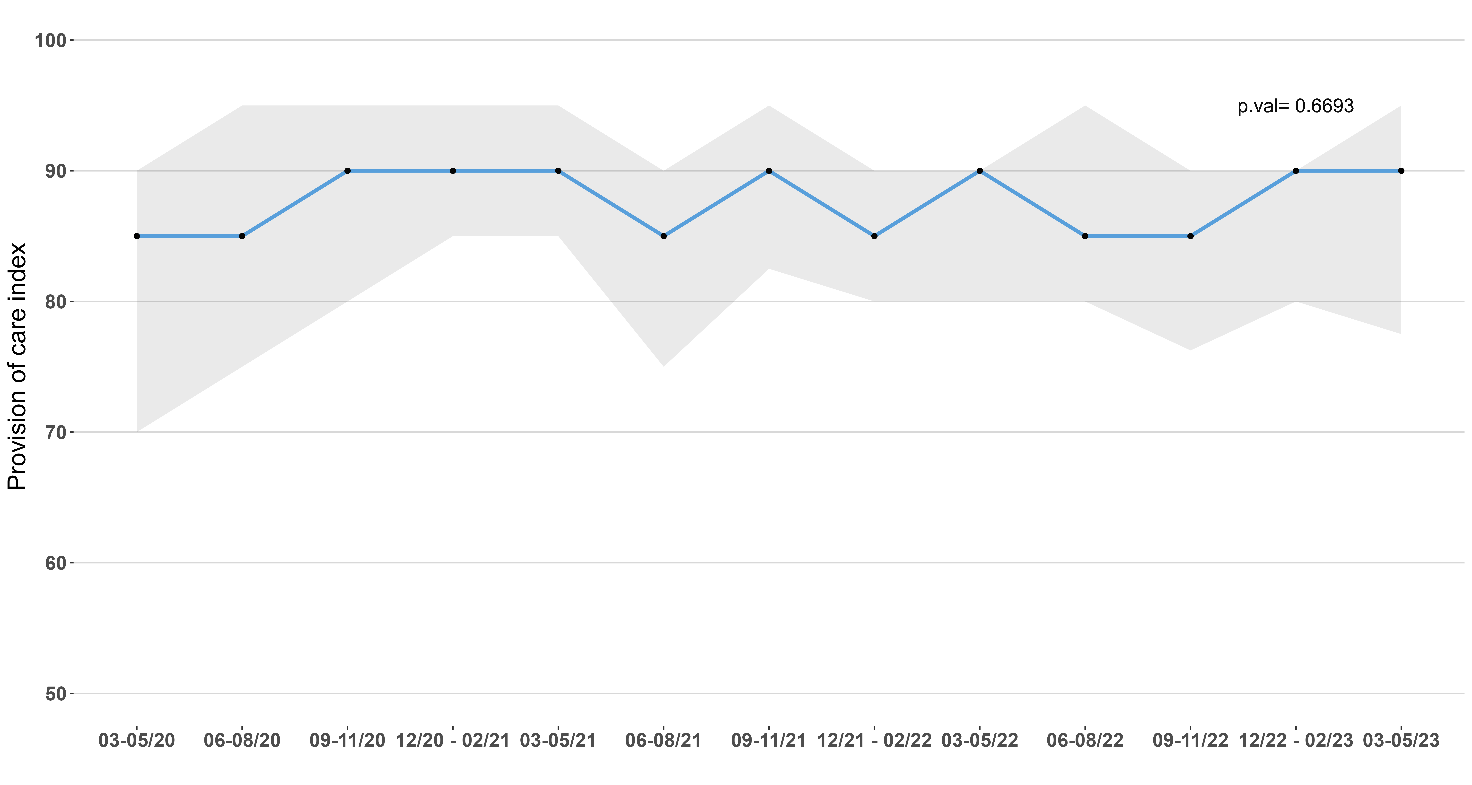

**Note**: the p-value is obtained with Mann‐Kendall test for monotonic trend (H0: no monotonic trend)

**Figure 2C: Lineplot showing the scores of Availability of physical and human resources index by trimester**

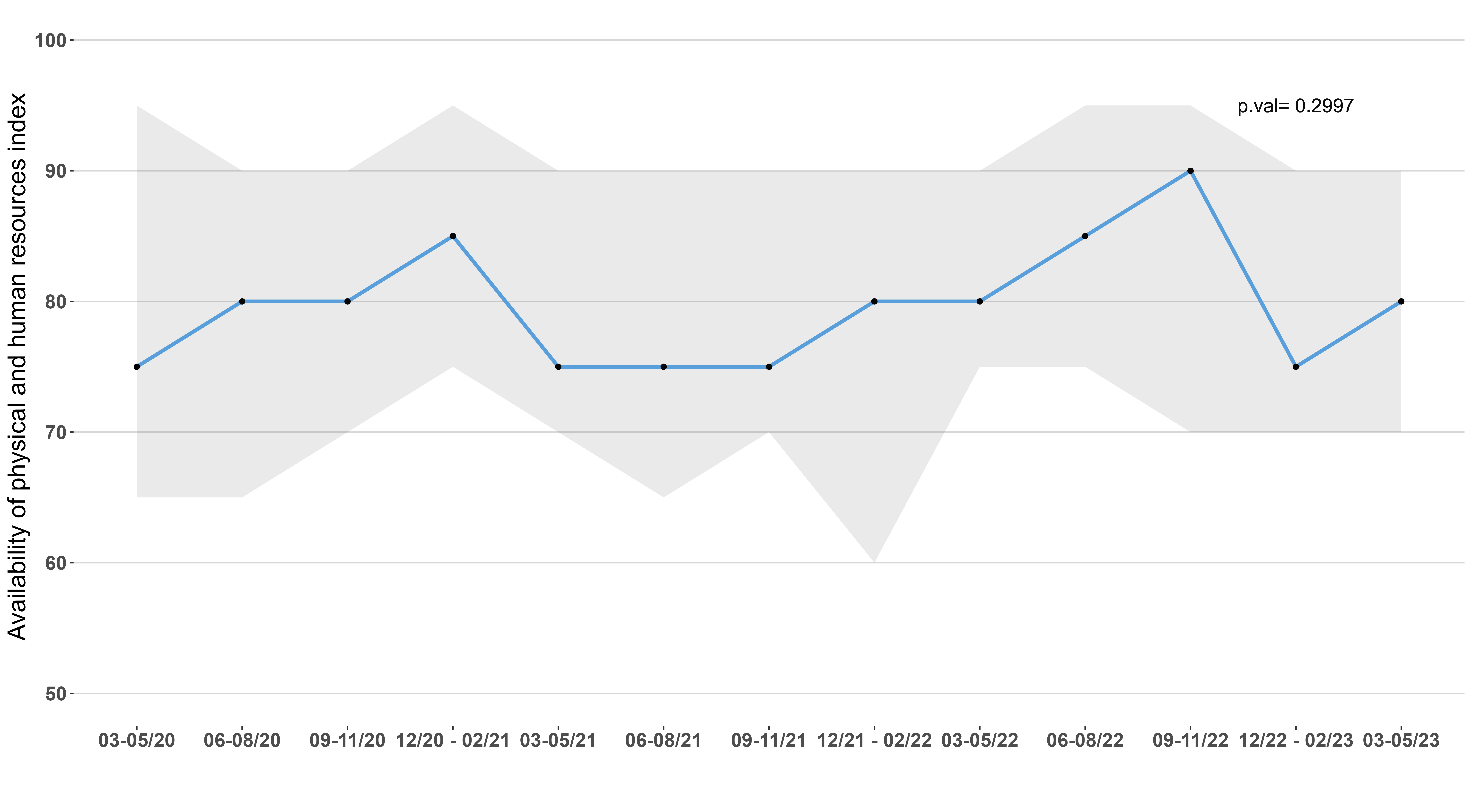

**Note**: the p-value is obtained with Mann‐Kendall test for monotonic trend (H0: no monotonic trend)
